# Cardiovascular risk in treated hypertensive patients with isolated diastolic hypertension and isolated low diastolic blood pressure

**DOI:** 10.1101/2023.08.11.23294003

**Authors:** Wei-Lun Chang, Ying-Fan Chen, Yu-Hsuan Lee, Ming-Neng Shiu, Po-Yin Chang, Chao-Yu Guo, Chi-Jung Huang, Chern-En Chiang, Chen-Huan Chen, the Taiwan Consortium of Hypertension-associated Cardiac Disease (TCHC) Study Group, Shao-Yuan Chuang, the Cardiovascular Diseases Risk Factor Two-township (CVDFACTs) Study Group, Hao-Min Cheng

## Abstract

**Background:** The prognosis of high or markedly low diastolic blood pressure (DBP) with normalized on-treatment systolic blood pressure (SBP) on major adverse cardiovascular events (MACE) is uncertain. This study examined whether isolated diastolic hypertension (IDH) and isolated low DBP (ILDBP) were associated with MACE in treated patients.

**Methods:** 7582 hypertensive patients with on-treatment SBP <130 mmHg from the Systolic Blood Pressure Intervention Trial (SPRINT) were categorized based on average DBP: <60 mmHg (n=1031; ILDBP), 60–79 mmHg (n=5432), ≥80 mmHg (n=1119; IDH). MACE risk was estimated using Cox proportional hazards models. The analysis was supplemented by a meta-analysis involving 10106 SPRINT and four cohort participants.

**Results:** Median age of the SPRINT participants was 67.0 years, and 64.9% were men. Over a median follow-up of 3.4 years, 512 patients developed MACE. The incidence of MACE was 3.9 cases per 100 person-years for ILDBP, 1.9 cases for DBP 60–79 mmHg, and 1.8 cases for IDH. ILDBP was associated with 1.32-fold increased MACE risk (hazard ratio [HR]: 1.32, 95% confidence interval [CI]: 1.05–1.66), whereas IDH was not (HR: 1.18, 95% CI: 0.87–1.59). There was no effect modification by age, sex, atherosclerotic cardiovascular disease risk, or cardiovascular disease history (all p-values for interaction >0.05). The meta-analysis indicated increased MACE risk in ILDBP (HR: 1.26, 95% CI: 1.05–1.52) compared with DBP 60–79 mmHg.

**Conclusions:** In treated patients with normalized SBP, excessively low DBP was associated with increased MACE risk, while IDH was not. Further research is required for ILDBP management.

## Introduction

Hypertension affects around 30% of adults globally [1] and is associated with increased cardiovascular risk [2-4]. The Systolic Blood Pressure Intervention Trial (SPRINT) revealed that intensive treatment targeting systolic BP (SBP) <120 mmHg was superior to standard treatment targeting SBP <140 mmHg in reducing cardiovascular events by 25% and all-cause mortality by 27% [5]. Consequently, the 2017 American College of Cardiology (ACC)/American Heart Association (AHA) guideline revised the BP treatment target to <130/80 mmHg [6].

For the patients on antihypertensive treatment, while the association between high SBP and cardiovascular risk is well-established, the potential harms posed by elevated diastolic BP (DBP) is still debated, especially when SBP achieves the target. This condition is known as isolated diastolic hypertension (IDH). Most studies examining the ACC/AHA definition of IDH have not found a correlation with cardiovascular disease [7-11], except in Asian populations and young adults [12-16]. The focus of these studies was primarily on treatment-naive individuals, leaving this correlation for antihypertensive drug-treated patients uncertain. Additionally, concerns also exist regarding the potential negative impact of excessive reduction of DBP, which could offset the cardiovascular benefits of reducing SBP.

The 2018 European Society of Cardiology (ESC)/European Society of Hypertension (ESH) guideline recommended a DBP target range of 70–80 mmHg for all risk levels [17], while the ACC/AHA guideline did not specify a lower limit of DBP for antihypertensive drug-treated patients [6]. However, these recommendations were primarily based on expert opinion rather than empirical evidence. It remains unknown whether high or extremely low DBP is associated with increased cardiovascular risk in antihypertensive drug-treated patients with normalized SBP.

To answer this question, we conducted a secondary analysis of SPRINT to determine if IDH and isolated low DBP (ILDBP) were associated with major adverse cardiovascular events (MACE) in patients with on-treatment SBP <130 mmHg. Additionally, we performed a meta-analysis of individual patient data (IPD) from SPRINT and four cohorts to supplement the initial analysis.

## Methods

### I. SPRINT Analysis

#### Data Sources and Study Participants

National Heart, Lung, and Blood Institute-managed Biologic Specimen and Data Repository Information Coordinating Center made anonymized SPRINT data available to the public for this study. The Institutional Review Board of Taipei Veterans General Hospital reviewed and approved the protocol for this investigation.

SPRINT [5] was an open-label randomized controlled trial, which enrolled 9361 participants between 2010–2013 aged ≥50 with SBP between 130–180 mmHg, increased cardiovascular risk, and no history of diabetes or stroke. The participants were randomly assigned to achieve a SBP goal of <120 mmHg (intensive treatment) or <140 mmHg (standard treatment). BP measurements and antihypertensive drug adjustments were scheduled monthly for the first three months, and then every three months thereafter. The trial intervention was terminated early in 2015 due to a significantly lower rate of the primary composite outcomes in the intensive treatment group than in the standard treatment group.

Our study utilized SPRINT data from each participant’s initial visit to their final visit. During the intervention and post-intervention periods, comprehensive participant information was meticulously recorded in the SPRINT data. For instance, demographic information was collected at the outset. During the follow-up period, clinical and laboratory data was collected at baseline and every three months. At each study visit, BP and antihypertensive drug use were recorded.

We selected participants from the SPRINT trial whose SBP readings during treatment were consistently <130 mmHg. We identified eligible participants by including those who had at least two consecutive SBP readings <130 mmHg, with Figure S1 providing further details on the subject selection process. To establish the starting point for follow-up, we designated the date of the first SBP reading <130 mmHg as the index date.

#### Cardiovascular Outcomes and Follow-up

The primary outcome was composed of MACE, including coronary heart disease (myocardial infarction and non-myocardial infarction acute coronary syndromes), stroke, heart failure, or cardiovascular death, occurring after the index date. Secondary outcomes were each individual component of the primary composite outcome.

The duration of follow-up for all study participants was calculated from the index date until the first occurrence of MACE or until the date of non-cardiovascular death or the last SPRINT visit.

#### Participant Classification Based on DBP Levels

Each participant’s DBP levels were calculated from all follow-up visits. For each participant, we calculated an average value by multiplying the DBP measured at each follow-up visit by the proportion of days between the date of BP measurement and the next measurement.

Using a multivariable adaptive regression spline analysis for MACE, the lower limit for average DBP during follow-up visits was established at 60 mmHg (Figure 1). Participants were divided into three groups based on their on-treatment DBP: <60 mmHg (ILDBP), 60–79 mmHg, and ≥80 mmHg (IDH).

**Figure 1.**
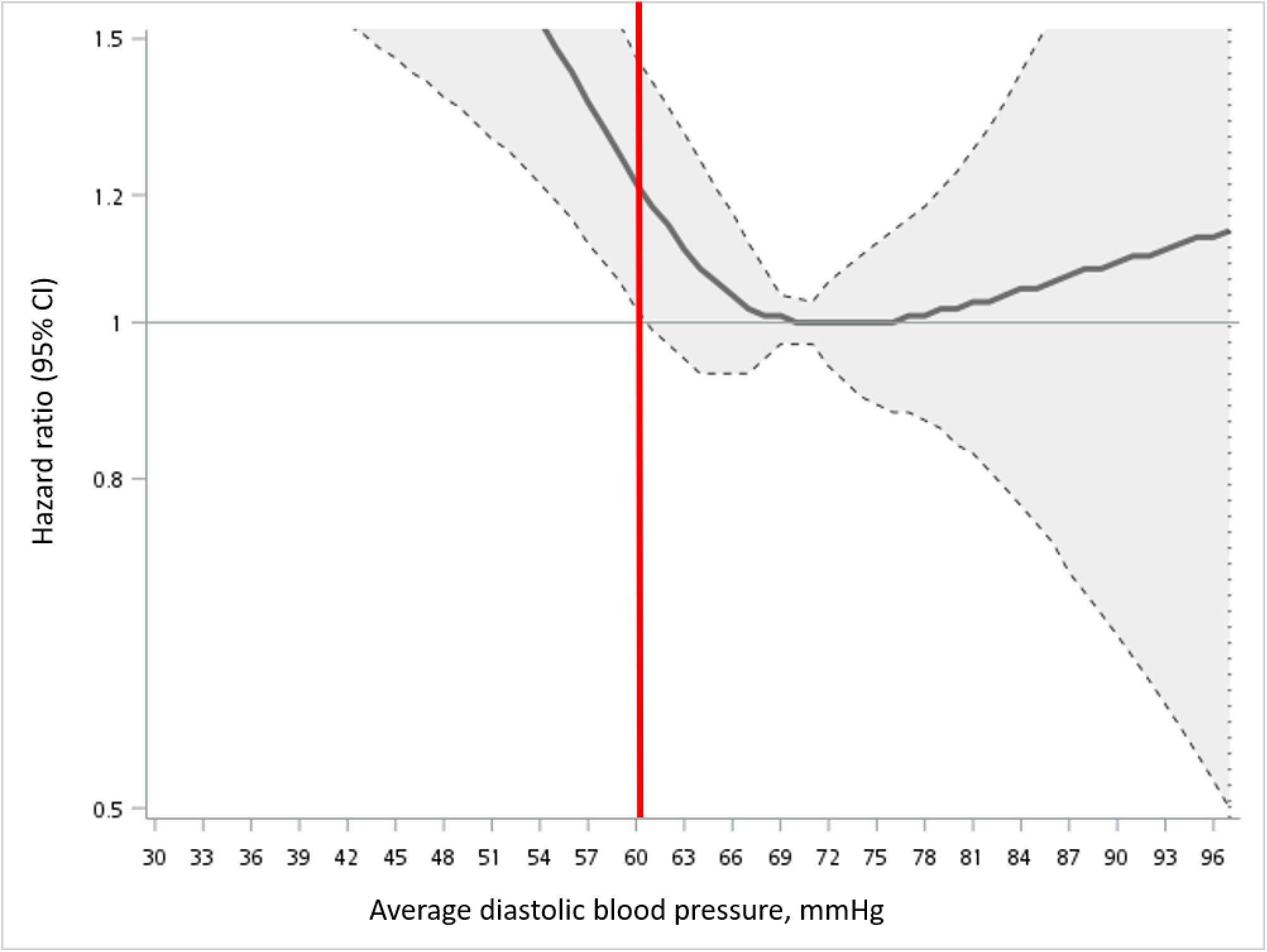
Cubic-spline curve for the association between average diastolic blood pressure and risk of major adverse cardiovascular events. The solid line indicates the hazard ratio for major adverse cardiovascular events and the dotted line indicates the 95% confidence interval. The hazard ratios were adjusted for age, sex, race, body mass index, total cholesterol, high-density lipoprotein cholesterol, plasma glucose, serum creatinine, smoking status, history of cardiovascular disease, number of antihypertensive drugs, risk of atherosclerotic cardiovascular disease, assigned treatment, and systolic blood pressure. The mean of average diastolic blood pressure (70 mmHg) was used as a reference. CI: confidence interval.

#### Statistical Analyses

Characteristics of study participants in each DBP group were summarized as means and standard deviations (continuous variables) or counts and percentages (categorical variables). We examined differences in characteristics between groups using analysis of variance tests and chi-squared tests. Time-varying Cox proportional hazards model was used to estimate the MACE risk, comparing IDH or ILDBP with DBP 60–79 mmHg, with adjustment for time-varying SBP over the follow-up, as well as time-invariant demographic characteristics, clinical and laboratory features, smoking status, history of cardiovascular disease, number of antihypertensive drugs, risk of atherosclerotic cardiovascular disease (ASCVD), and assigned treatment. In addition to the analysis of three DBP groups, the average DBP was analyzed using deciles as a categorical variable. Due to the nonlinear relationship between average DBP and cardiovascular disease, we used the deviation from the mean coding to estimate the risk in the deciles of average DBP [18]. SBP, age, clinical and laboratory characteristics, number of antihypertensive drugs, and ASCVD risk were treated as continuous variables, whereas the remaining variables were treated as categorical variables. To reduce potential bias due to missing data, missing values on covariates were estimated using multiple imputations. Results of Cox proportional hazards models were presented as hazard ratios (HRs) and 95% confidence intervals (CIs).

Subgroup analyses were performed to explore whether the HR for MACE in relation to the DBP groups differed between age (50–64 years and ≥65 years), sex (male and female), ASCVD risk (<10% and ≥10%), and history of cardiovascular disease (yes and no).

Sensitivity analyses were performed to assess the robustness of results. In the initial analysis, the Cox proportional hazards models were repeated after additional adjustment for duration between the start of SPRINT follow-up and index date. In the second analysis, we used 85 mmHg or 90 mmHg as a cutoff determining the IDH group to examine if the risk for IDH would vary with different thresholds. In the third analysis, given the high degree of correlation between SBP and DBP, we decorrelated the two BP indices by regressing SBP on DBP and utilizing the SBP residual to estimate the HRs. Following this, a heatmap was created to illustrate the contribution of the DBP and SBP in their associations with MACE.

A two-tailed p-value of <0.05 was considered to indicate statistical significance. Data processing and statistical analyses were performed using SAS (Version 9.4).

### II. IPD Meta-analysis

The supplementary analysis was carried out with a two-stage IPD meta-analysis of SRPINT and four large cohorts - Coronary Artery Risk Development in Young Adults (CARDIA) [18], Cardiovascular Diseases Risk Factor two-Township Study (CVDFACTs) [19, 20], Jackson Heart Study (JHS) [21], and Taiwan Consortium of Hypertension-associated Cardiac disease study (TCHC) [22]. All studies have collected BP data of a research-grade quality. The protocol for this study was approved by the Institutional Review Board of Taipei Veterans General Hospital.

Table S1 provides information on the recruitment in detail. For each cohort, we identified individuals treated with antihypertensive medications whose baseline SBP was <130 mmHg. Following the same cutoffs, 60 mmHg and 80 mmHg, as in the SPRINT analysis, we used office BP at baseline, a measurement method used in all cohorts, to classify the three DBP groups. All participants were followed from baseline until the first occurrence of MACE or until the date of non-cardiovascular death or until the end of the study. Detailed information on the methods to measure office BP and the definition of MACE is presented in Table S2 and Table S3, respectively.

We conducted the IPD meta-analysis with two stage approaches. First, we estimated the MACE risk using multivariable Cox proportional hazards models, comparing IDH or ILDBP with DBP 60–79 mmHg for each study. Except for the use of antihypertensive drugs, the definitions of covariates were identical to those used in the SPRINT analysis. When analyzing the JHS and TCHC data, we defined the number of antihypertensive drugs based on the intake of angiotensin-converting enzyme inhibitor, angiotensin receptor blocker, beta-blocker, calcium channel blocker, and diuretic. However, the information on the drug use is not available in the CARDIA and CVDFACTs data. Analyses were performed after multiple imputations for missing data. Next, we combined the study-specific estimates of adjusted HR using a random-effects model and subsequently stratified the results according to age (<50 years, 50–64 years, and ≥65 years), sex (male and female), ASCVD risk (<10% and ≥10%), and race (Asian and non-Asian). As a cutoff for determining IDH, 130/80 mmHg or 140/90 mmHg was utilized in sensitivity analyses. Heterogeneity for pooled outcomes was assessed with the I² statistics and p-values for chi-squared tests. The statistical analyses were performed using Review Manager (Version 5.4).

## Results

### I. SPRINT Analysis

#### Characteristics of Study Participants

A total of 7582 SPRINT participants aged ≥50 with at least two consecutive readings of SBP <130 mmHg were identified for analysis (Figure S1). These participants’ median age was 67.0 years, and 64.9% of them were men. The participants were divided into three groups according to their average DBP levels: <60 mmHg (n=1031), 60–79 mmHg (n=5432), and ≥80 mmHg (n=1119). Compared to the other two DBP groups, ILDBP participants were more likely to be older, never-smokers, have a lower body mass index and total cholesterol, and have greater risk of ASCVD. The participants with IDH, who were more likely to be younger and active smokers, had a greater body mass index and total cholesterol but decreased risk of ASCVD (Table 1).

**Table 1.** Characteristics of the study participants by diastolic blood pressure group.

|  | DBP <60 | DBP 60–79 | DBP ≥80 |  |
| --- | --- | --- | --- | --- |
|  | mmHg | mmHg | mmHg |  |
| Characteristics | (n=1031) | (n=5432) | (n=1119) | P value |
| Age, years | 76.6 ±7.4 | 68.0 ±8.9 | 61.5 ±7.0 | <.0001 |
| Age group, n (%) |  |  |  | <.0001 |
| 50–64 years | 79 (7.7%) | 2112 (38.9%) | 791 (70.7%) |  |
| ≥65 years | 952 (92.3%) | 3320 (61.1%) | 328 (29.3%) |  |
| Male, n (%) | 673 (65.3%) | 3494 (64.3%) | 751 (67.1%) | 0.1959 |
| Race, n (%) |  |  |  | <.0001 |
| White | 730 (70.8%) | 3237 (59.6%) | 502 (44.9%) |  |
| Black | 186 (18.0%) | 1535 (28.3%) | 515 (46.0%) |  |
| Hispanic | 90 (8.7%) | 553 (10.2%) | 90 (8.0%) |  |
| Other | 25 (2.4%) | 107 (2.0%) | 12 (1.1%) |  |
| SBP at the index date,<br>mmHg | 119.6 ±7.6 | 119.2 ±7.6 | 120.4 ±7.5 | <.0001 |
| DBP at the index date,<br>mmHg | 56.0 ±7.0 | 68.7 ±8.0 | 78.1 ±7.1 | <.0001 |
| PP at the index date,<br>mmHg | 63.7 ±8.3 | 50.6 ±8.3 | 42.2 ±6.6 | <.0001 |
| BMI, kg/m<sup>2</sup> | 28.0 ±5.4 | 30.1 ±5.7 | 31.5 ±6.0 | <.0001 |
| Total cholesterol, mg/dL | 178.0 ±37.7 | 188.4 ±41.4 | 196.5 ±41.5 | <.0001 |
| HDL cholesterol, mg/dL | 54.1 ±14.4 | 52.4 ±14.0 | 51.4 ±13.7 | 0.0015 |
| Plasma glucose, mg/dL | 98.6 ±11.3 | 99.1 ±14.9 | 98.4 ±17.2 | 0.5338 |
| Serum creatinine, mg/dL | 1.2 ±0.5 | 1.1 ±0.4 | 1.1 ±0.4 | <.0001 |
| Smoking status, n (%) |  |  |  | <.0001 |
| Current | 65 (6.3%) | 644 (11.9%) | 217 (19.4%) |  |
| Non-current | 965 (93.6%) | 4785 (88.1%) | 900 (80.4%) |  |
| Missing data | 1 (0.1%) | 3 (0.1%) | 2 (0.2%) |  |
| DM history, n (%) | 0 (0.0%) | 10 (0.2%) | 0 (0.0%) | 0.2113 |
| CVD history, n (%) | 313 (30.4%) | 1127 (20.8%) | 150 (13.4%) | <.0001 |
| No. of antihypertensive drugs | 2.8 ±1.2 | 2.3 ±1.1 | 2.0 ±1.1 | <.0001 |
| ASCVD risk, % | 26.9 ±12.8 | 17.1 ±10.3 | 12.5 ±6.0 | <.0001 |
| ASCVD risk group, n (%) |  |  |  | <.0001 |
| <10% | 38 (3.7%) | 929 (17.1%) | 242 (21.6%) |  |
| ≥10% | 607 (58.9%) | 2761 (50.8%) | 462 (41.3%) |  |
| Missing data | 386 (37.4%) | 1742 (32.1%) | 415 (37.1%) |  |
| Intensive treatment, n (%) | 810 (78.6%) | 3398 (62.6%) | 208 (18.6%) | <.0001 |
| Duration between the start of SPRINT follow-up and index date, days | 179.8 ±287.4 | 166.6 ±278.1 | 208.9 ±308.2 | <.0001 |

Data are expressed as mean ±SD or n (%).
DBP: diastolic blood pressure; SBP: systolic blood pressure; PP: pulse pressure; BMI: body mass index; HDL: high-density lipoprotein; DM: diabetes mellitus; CVD: cardiovascular
disease; ASCVD: atherosclerotic cardiovascular disease; SPRINT: Systolic Blood Pressure Intervention Trial.

#### Overall Analyses

During a median (interquartile range) follow-up of 3.4 (2.7–4.0) years, 512 participants developed MACE. Figure S2 illustrates the HRs and 95% confidence intervals (CIs) for MACE according to deciles of average DBP, with adjustment for covariates. Only in the lowest decile group (average DBP <58 mmHg) was the risk significantly higher than the population average risk (adjusted HR [aHR]: 1.40, 95% confidence interval [CI]: 1.10–1.77).

When participants were divided into three DBP groups, the incidence of MACE per 100 person-years were 3.9 for ILDBP, 1.9 for DBP 60–79 mmHg, and 1.8 for IDH. Compared with DBP 60–79 mmHg, ILDBP was associated with 1.32-fold increased risk of MACE (aHR: 1.32, 95% CI: 1.05–1.66), whereas IDH was not (aHR: 1.18, 95% CI: 0.87–1.59) (Table 2). ILDBP was associated with increased risk for all MACE types, with coronary heart disease having the highest risk (aHR: 1.37, 95% CI: 1.28–1.46) (Table 2).

**Table 2.** Event rates and hazard ratios of major adverse cardiovascular events by diastolic blood pressure group.

|  | No. of | Event rate per | Unadjusted |  | Adjusted<sup>a</sup> |  | Adjusted<sup>b</sup> |  |
| --- | --- | --- | --- | --- | --- | --- | --- | --- |
| Outcome | events (%) | 100 person-years | HR (95% CI) | P value | HR (95% CI) | P value | HR (95% CI) | P value |
| *Primary outcome* |  |  |  |  |  |  |  |  |
| Major adverse cardiovascular events |  |  |  |  |  |  |  |  |
| DBP 60–79 mmHg | 328 (6.0%) | 1.9 | Reference |  | Reference |  | Reference |  |
| DBP ≥80 mmHg | 61 (5.5%) | 1.8 | 0.95 (0.84-1.07) | 0.4137 | 1.18 (0.87-1.59) | 0.2986 | 1.19 (0.88-1.61) | 0.2616 |
| DBP <60 mmHg | 123 (11.9%) | 3.9 | 2.12 (1.93-2.32) | <.0001 | 1.32 (1.05-1.66) | 0.0188 | 1.32 (1.05-1.67) | 0.0186 |
| *Secondary outcome* |  |  |  |  |  |  |  |  |
| Coronary heart disease |  |  |  |  |  |  |  |  |
| DBP 60–79 mmHg | 159 (2.9%) | 0.9 | Reference |  | Reference |  | Reference |  |
| DBP ≥80 mmHg | 29 (2.6%) | 0.8 | 0.93 (0.78-1.11) | 0.4305 | 1.06 (0.97-1.16) | 0.1919 | 1.04 (0.95-1.14) | 0.3712 |
| DBP <60 mmHg | 58 (5.6%) | 1.8 | 2.07 (1.81-2.36) | <.0001 | 1.37 (1.28-1.46) | <.0001 | 1.38 (1.29-1.49) | <.0001 |
| Stroke |  |  |  |  |  |  |  |  |
| DBP 60–79 mmHg | 62 (1.1%) | 0.3 | Reference |  | Reference |  | Reference |  |
| DBP ≥80 mmHg | 8 (0.7%) | 0.2 | 0.66 (0.48-0.92) | 0.0134 | 1.09 (1.00-1.18) | 0.0443 | 1.06 (0.97-1.14) | 0.1958 |
| DBP <60 mmHg | 24 (2.3%) | 0.7 | 2.14 (1.73-2.64) | <.0001 | 1.33 (1.26-1.42) | <.0001 | 1.35 (1.27-1.44) | <.0001 |
| Heart failure |  |  |  |  |  |  |  |  |
| DBP 60–79 mmHg | 90 (1.7%) | 0.5 | Reference |  | Reference |  | Reference |  |
| DBP ≥80 mmHg | 15 (1.3%) | 0.4 | 0.90 (0.70-1.14) | 0.3749 | 1.01 (0.93-1.10) | 0.8287 | 0.99 (0.91-1.08) | 0.8055 |
| DBP <60 mmHg | 45 (4.4%) | 1.4 | 3.10 (2.65-3.63) | <.0001 | 1.30 (1.22-1.38) | <.0001 | 1.31 (1.23-1.40) | <.0001 |
| Cardiovascular death |  |  |  |  |  |  |  |  |
| DBP 60–79 mmHg | 54 (1.0%) | 0.3 | Reference |  | Reference |  | Reference |  |
| DBP ≥80 mmHg | 13 (1.2%) | 0.4 | 1.28 (0.98-1.68) | 0.0744 | 1.04 (0.96-1.12) | 0.3682 | 1.02 (0.94-1.10) | 0.6991 |
| DBP <60 mmHg | 26 (2.5%) | 0.8 | 2.85 (2.31-3.50) | <.0001 | 1.31 (1.23-1.39) | <.0001 | 1.32 (1.25-1.41) | <.0001 |

DBP: diastolic blood pressure; HR: hazard ratio; CI: confidence interval.

#### Subgroup Analyses

There was no effect modification by age group, sex, ASCVD risk, or cardiovascular disease history. The aHR for MACE in ILDBP was similar between participants aged 50–64 and those aged ≥65 years (p-value for interaction: 0.7994). The same was observed for IDH (p-value for interaction: 0.0550) (Figure 2). The aHR of MACE in ILDBP and IDH were comparable in male and female participants (p-value for interaction for ILDBP: 0.5491; for IDH: 0.4952), as well as in participants with low and high ASCVD risk (p-value for interaction for ILDBP: 0.9785; for IDH: 0.1381) (Figure 2). The correlation between ILDBP or IDH and MACE remained consistent for both patients with and without a cardiovascular disease history (p-value for interaction for ILDBP: 0.8214; for IDH: 0.4322) (Figure 2).

**Figure 2.**
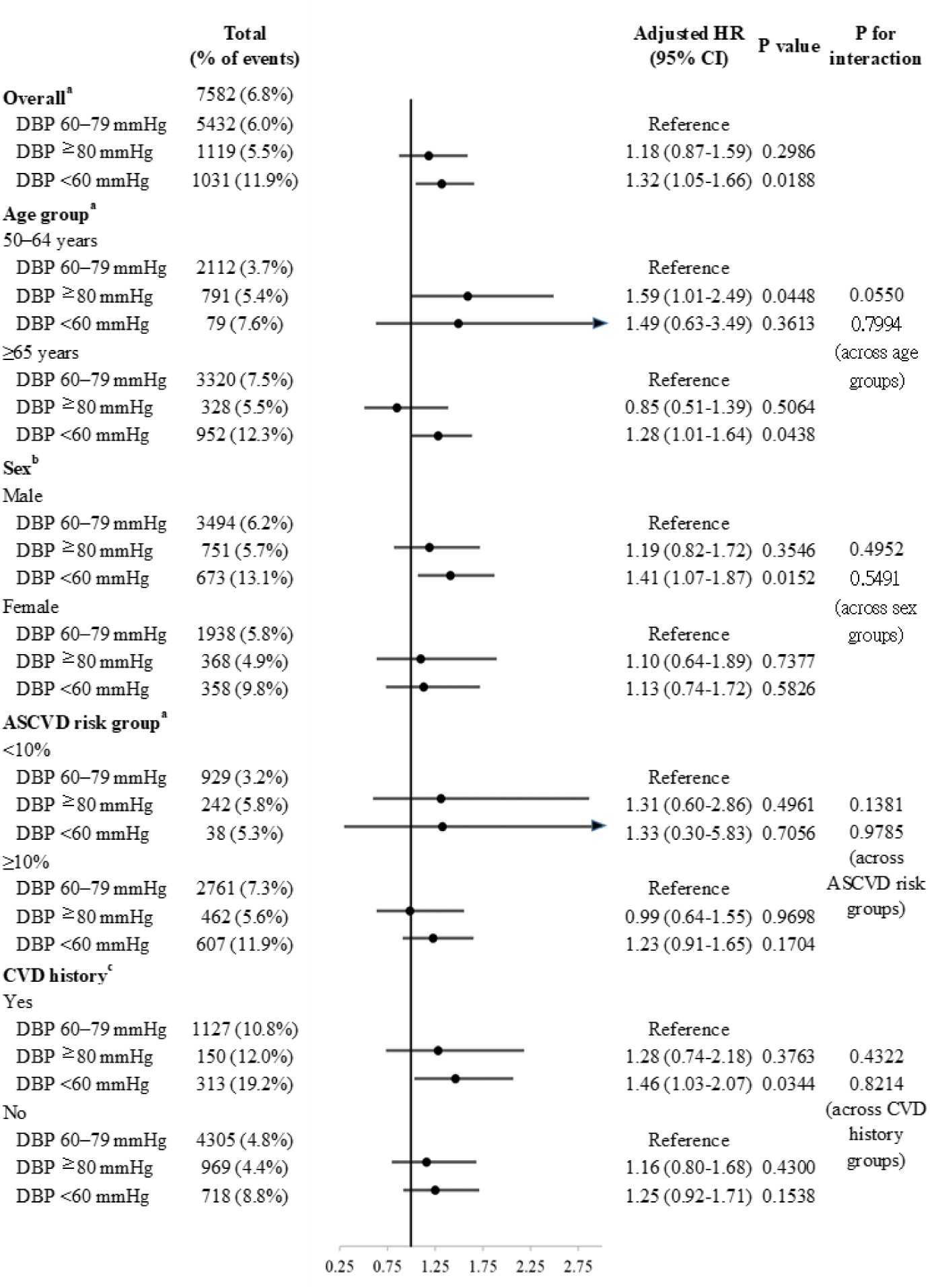
Subgroup analyses of hazard ratios of major adverse cardiovascular events according to age, sex, risk of atherosclerotic cardiovascular disease, and history of cardiovascular disease. a: Adjusted for age, sex, race, body mass index, total cholesterol, high-density lipoprotein cholesterol, plasma glucose, serum creatinine, smoking status, history of cardiovascular disease, number of antihypertensive drugs, risk of atherosclerotic cardiovascular disease, assigned treatment, and systolic blood pressure. b: Adjusted for age, race, body mass index, total cholesterol, high-density lipoprotein cholesterol, plasma glucose, serum creatinine, smoking status, history of cardiovascular disease, number of antihypertensive drugs, risk of atherosclerotic cardiovascular disease, assigned treatment, and systolic blood pressure. c: Adjusted for age, sex, race, body mass index, total cholesterol, high-density lipoprotein cholesterol, plasma glucose, serum creatinine, smoking status, number of antihypertensive drugs, risk of atherosclerotic cardiovascular disease, assigned treatment, and systolic blood pressure. DBP: diastolic blood pressure; ASCVD: atherosclerotic cardiovascular disease; HR: hazard ratio; CI: confidence interval.

#### Sensitivity Analyses

The results of the first sensitivity analysis, with further adjustment for the length of time between the start of SPRINT follow-up and the index date, were comparable to those found in the primary analysis (Table 2). The second sensitivity analysis applying other IDH cutoffs gave comparable results to the initial analysis (Table S4). The third sensitivity analysis using the residual method revealed an association between ILDBP (aHR: 1.34, 95% CI: 1.07–1.69) or IDH (aHR: 1.15, 95% CI: 0.75–1.56) and MACE comparable to the primary analysis. The heatmap revealed that along the vertical axis, 3-year risk of MACE were significantly associated with DBP levels (p-value: 0.0068) (Figure S3).

### II. IPD Meta-analysis

A total of 10106 participants were extracted from the SPRINT and four cohorts. A summary of the participant characteristics analyzed in this study is reported in Table S5.

Participants were divided into three DBP groups, similar to the SPRINT analysis, the results suggested significantly higher risk for MACE in ILDBP (aHR: 1.26, 95% CI: 1.05– 1.52), compared with DBP 60–79 mmHg. Also, no significant association was found between IDH and MACE (aHR: 0.88, 95% CI: 0.57–1.35) (Figure 3 and Figure S4).

**Figure 3.**
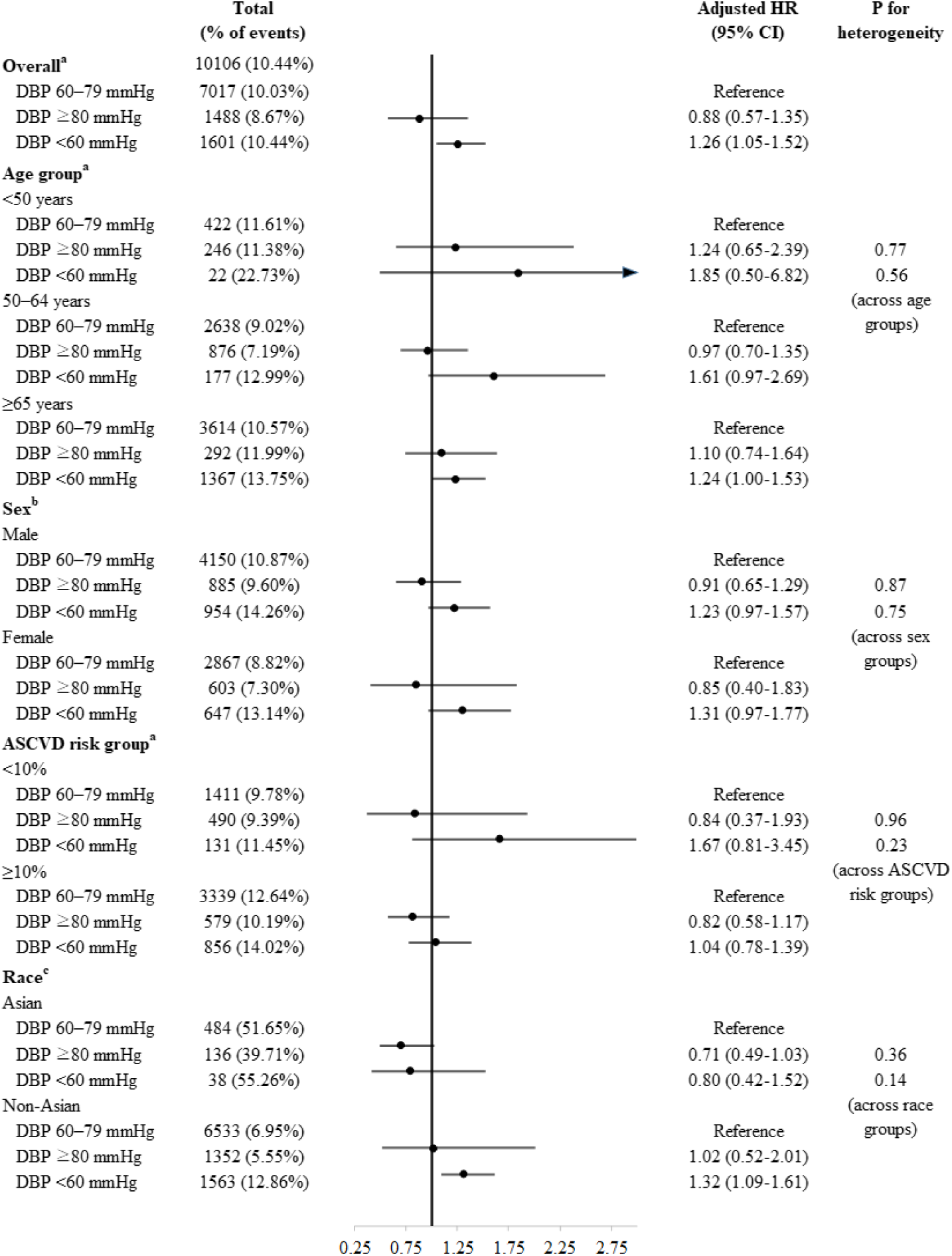
Overall and subgroup results of the individual participant meta-analyses. a: Adjusted for age, sex, race, body mass index, total cholesterol, high-density lipoprotein cholesterol, plasma glucose, serum creatinine, smoking status, history of cardiovascular disease, number of antihypertensive drugs, risk of atherosclerotic cardiovascular disease, assigned treatment (in SPRINT), and systolic blood pressure. b: Adjusted for age, race, body mass index, total cholesterol, high-density lipoprotein cholesterol, plasma glucose, serum creatinine, smoking status, history of cardiovascular disease, number of antihypertensive drugs, risk of atherosclerotic cardiovascular disease, assigned treatment (in SPRINT), and systolic blood pressure. c: Adjusted for age, sex, body mass index, total cholesterol, high-density lipoprotein cholesterol, plasma glucose, serum creatinine, smoking status, history of cardiovascular disease, number of antihypertensive drugs, risk of atherosclerotic cardiovascular disease, assigned treatment (in SPRINT), and systolic blood pressure. DBP: diastolic blood pressure; ASCVD: atherosclerotic cardiovascular disease; HR: hazard ratio; CI: confidence interval.

The heterogeneity tests showed that the differences in adjusted hazard ratios across age groups (p-value: 0.77 and 0.56), sexes (p-value: 0.87 and 0.75), ASCVD risk groups (p-value: 0.96 and 0.23), and race (p-value: 0.36 and 0.14) were not statistically significant (Figure 3).

The sensitivity analysis for the IPD meta-analysis, which utilized different IDH cutoffs, showed that neither IDH nor ILDBP was associated with increased MACE risk (Table S6).

## Discussion

This study investigated the association between IDH and ILDBP with MACE in antihypertensive-treated patients with normalized SBP. The analysis of SPRINT and IPD meta-analysis indicated that individuals with ILDBP had higher risk of MACE compared to those with normalized SBP and DBP between 60–79 mmHg, while IDH did not. These findings were consistent across different age groups, sexes, atherosclerotic cardiovascular disease risk, and cardiovascular disease history.

In our SPRINT analysis, patients classified as having IDH according to the 2017 ACC/AHA guidelines showed similar risk MACE compared to those with DBP between 60– 79 mmHg. However, previous studies examining the cardiovascular risk of IDH have produced conflicting results, which may be due to variations in study comparators or the low prevalence of IDH among older individuals. For example, Yano et al. found discrepant outcomes in different age groups, suggesting the heterogeneity of IDH patients and the need for extensive phenotyping to identify those at higher cardiovascular risk [23]. This highlights the need for extensive phenotyping to identify IDH patients at higher risk for cardiovascular disease, as they are likely a heterogeneous group. Jacobsen et al. also noted that individuals with stage 1 IDH did not consistently exhibit elevated cardiovascular disease risk compared to those with normal BP [24]. Therefore, these patients do not require drug treatment and should instead undergo lifestyle changes, interval BP checks, and only considered for drug treatment if systolic hypertension or stage 2 IDH develops. Some meta-analyses and the International Database of Ambulatory blood pressure in relation to Cardiovascular Outcome (IDACO) trial have found increased cardiovascular risk in young patients with IDH, but not overall mortality [23, 25, 26], which contrasts with previous studies. The physiological explanation for this discrepancy remains unclear.

Additionally, a study using a DBP threshold of 90 mmHg did not show a significant increase in the risk of MACE in IDH based on ESC/ESH criteria [17]. Several post hoc studies that found a linear association between DBP level and CVD risk seem to challenge this finding [27]. The increased risk observed in ILDBP may be attributed to the close association between high DBP and elevated SBP, which is known to be linked with cardiovascular risk. Excluding participants with SBP above 130 mmHg may decrease the likelihood of DBP elevation. Furthermore, our study revealed elevated risk in the ILDBP group when using a cutoff of 130/80 mmHg, indicating that the ACC/AHA classification for hypertension accurately assesses this unique phenotype. Notably, our research demonstrated that lower DBP levels were associated with worse clinical outcomes, with ILDBP posing higher risk compared to DBP in the range of 60–79 mmHg. This J-curve pattern has been observed in previous studies, including Khan et al. [28]. Their analysis of SPRINT data revealed that DBP <55 mmHg was associated with 25% higher risk compared to 70 mmHg. This may be due to intensive treatment affecting coronary artery blood flow autoregulation and increasing the risk of ischemia. Lee et al. found a higher risk of adverse outcomes in SPRINT participants experiencing diastolic hypotension with a baseline DBP ≥65 mmHg [29]. Further randomized controlled trials are needed to clarify this issue.

ILDBP, similar to isolated systolic hypertension, represents a form of vascular aging characterized by increased arterial stiffness and pulse pressure. Given the well-established role of SBP as a major cardiovascular risk factor requiring intensive treatment, it is crucial for patients with ILDBP to maintain SBP levels within the target range. Further research is needed to develop effective treatment strategies for ILDBP.

Our study has notable merits. First, we included a significant number of hypertensive patients from diverse populations by selecting participants from SPRINT and large multinational cohorts. Second, in the secondary analysis of SPRINT, we employed multiple BP measures for the DBP categorization, reducing the potential for misclassification bias of DBP status. Additionally, we considered repeated SBP readings as a time-varying variable to account for their contribution to cardiovascular events.

Yet, our study has several limitations. First, since we assessed secondary data from a randomized controlled trial, unmeasured confounding cannot be ruled out completely. Second, the absence of biomarkers of vascular aging hinders our understanding of the underlying mechanism between treated low DBP and cardiovascular risk. Although our findings suggest the existence of a new hypertension phenotype, ILDBP, confirming the causal relationship between on-treatment low DBP and increased risk requires randomized controlled trials.

In conclusion, in antihypertensive-treated patients with SBP of less than 130 mmHg, excessively low DBP was associated increased MACE risk. However, there is no evidence of higher MACE risk for IDH. Further study is needed to develop appropriate treatments for ILDBP.

## Data Availability

All data produced in the present work are contained in the manuscript.

## Acknowledgements

We gratefully acknowledge the participants involved in the Systolic Blood Pressure Intervention Trial (SPRINT) and the four cohort participants included in the meta-analysis. We also express gratitude to the investigators who took part in data collection and management.

## Source of Funding

None.

## Disclosures

All authors have nothing to disclose or conflicts of interest related to this manuscript.

## Notes

### Competing Interest Statement

The authors have declared no competing interest.

### Funding Statement

No funding was received for this work.

### Author Declarations

The Institutional Review Board of Taipei Veterans General Hospital reviewed and approved the protocol for this investigation.

